# Serological study of CoronaVac vaccine and booster doses in Chile: immunogenicity and persistence of anti-SARS-CoV-2 S antibodies

**DOI:** 10.1101/2022.01.14.22269289

**Authors:** Leonardo Vargas, Nicolás Valdivieso, Fabián Tempio, Valeska Simon, Daniela Sauma, Lucía Valenzuela, Caroll Beltrán, Loriana Castillo-Delgado, Ximena Contreras-Benavides, Mónica L. Acevedo, Fernando Valiente-Echeverría, Ricardo Soto-Rifo, Rafael I. Gonzalez, Mercedes Lopez, Fabiola Osorio, María Rosa Bono

## Abstract

**Background:** Chile was severely affected by COVID19 outbreaks but was also one of the first countries to start a nationwide program to vaccinate against the disease. Furthermore, Chile became one of the fastest countries to inoculate a high percentage of the target population and implemented homologous and heterologous booster schemes in late 2021 to prevent potential immunological waning. The aim of this study is to compare the immunogenicity and time course of the humoral response elicited by the CoronaVac vaccine in combination with homologous versus heterologous boosters.

**Methods and Findings:** We compared the immunogenicity of two doses of CoronaVac and BNT162b2 vaccines and studied the effect of different booster regimes in the Chilean population. Our results demonstrate that a two-dose vaccination scheme with CoronaVac induces lower levels of anti-SARS-CoV-2 S antibodies than BNT162b2 in a broad age range. Furthermore, antibody production declines with time in individuals vaccinated with CoronaVac and less noticeably, with BNT162b2. Remarkably, analysis of booster schemes revealed that individuals vaccinated with two doses of CoronaVac generate immunological memory against the SARS-CoV-2 ancestral strain, which can be re-activated with homologous or heterologous (BNT162b2 and ChAdOx1) boosters. Nevertheless, the magnitude of the antibody response with the heterologous booster regime was considerably higher and persistent (over 100 days) than the responses induced by the homologous scheme.

**Conclusions:** Two doses of CoronaVac induces antibody titers against the SARS-CoV-2 ancestral strain which are lower in magnitude than those induced by the BNT162b2 vaccine. However, the response induced by CoronaVac can be greatly potentiated with a heterologous booster scheme with BNT162b2 or ChAdOx1 vaccines. Furthermore, the heterologous booster regimes induce a durable antibody response which does not show signs of decay 3 months after the booster dose.

## INTRODUCTION

Chile is one of the several countries severely threatened by the COVID-19 pandemic in 2020, but that had prompt access to vaccines for a large number of individuals since early 2021. The first SARS-CoV-2 vaccine authorized in Chile for emergency use by the Health Ministry (MINSAL) was the Pfizer-BioNTech vaccine (BNT162b2) on December 16, 2020, and Sinovac’s CoronaVac vaccine on January 20, 2021 (Institute of Public Health, ISP). World Health Organization (WHO) listed CoronaVac for emergency use on June 1, 2021 (https://covid19.trackvaccines.org/agency/who/), which is currently administered in 48 countries (https://covid19.trackvaccines.org/vaccines/7/). In Chile, vaccination with CoronaVac began on February 1, 2021, with people over 55 years old, people with specific pathologies, and essential services personnel. Progressively, the vaccination scheme extended to younger people (target population over 18 years old: 15,200,840). In this first phase of vaccination, the CoronaVac vaccine was predominantly used across the population. Real-world data indicated that the two-dose vaccination scheme with CoronaVac in Chile showed a 65.9% vaccine effectiveness, 90.3% for prevention of ICU admission, and 86.3% for prevention of COVID-19 related death (1). To date, more than 86,8% of the Chilean population received their complete vaccination schedule with any available vaccines (DEIS/MINSAL), and about 77% of the target population received CoronaVac (Minsal / Deis).

However, around mid-2021, immunological studies reported a decline of antibody levels in vaccinated individuals. These studies predicted a reduction in antibody titers directed against SARS-CoV-2 over time, highlighting the requirement of an additional immunization (2) (3). In this context, a group of countries, including Israel (4), and Chile authorized a booster vaccine dose. On August 11, 2021, the vaccination with booster doses began for people who had received two doses of Coronavac in Chile. Interestingly, Chile implemented a heterologous booster schedule for most individuals including BNT162b2 and the ChAdOx1 vaccine from AstraZeneca as the most used boosters. These schemes offer an important opportunity to assess the magnitude of the immunological response to homologous and heterologous boosters schedules within the same population. Furthermore, this issue is relevant considering that immunological studies of heterologous booster schedules using CoronaVac as the first immunization vaccine have not been extensively documented.

This study describes the production of IgG antibodies directed against the ancestral SARS-CoV-2 S protein induced by the two-dose scheme of the Coronavac vaccine in a Health Service of the Hospital La Florida, Santiago. Our data shows that detectable levels of specific antibodies appear in most vaccinated individuals. By comparing the humoral responses to CoronaVac and BNT162b2 vaccines over time, we found that the antibody production elicited by CoronaVac declined six months after vaccination, whereas people vaccinated with two doses of BNT162b2 maintained a noticeably higher level of antibodies over time. Next, we analyzed the impact of the booster doses of CoronaVac, BNT162b2, or ChAdOx1 vaccines, administered to individuals vaccinated with the two-dose scheme with CoronaVac six months earlier. Our data show that the three types of boosters produce a noticeable increase in anti-spike IgG antibody production twenty days after the booster administration, which was more strongly noticed in individuals vaccinated with the heterologous booster regime. Antibody responses measured 100 days after the booster dose revealed that the heterologous regime induced higher and persistent anti-SARS-CoV-2 S antibodies compared to the homologous regime.

In summary, our results show that the CoronaVac vaccine produces memory against the SARS-CoV-2 that can be greatly potentiated with a heterologous booster strategy. Moreover, the persistent antibody titers obtained using the heterologous booster strategy may allow to space subsequent booster doses in the population. Furthermore, these data suggests that Chile’s vaccination scheme has been efficient in avoiding contagion with the Delta variant, as predicted by data derived from the epidemic in Chile.

## MATERIALS AND METHODS

### Ethics Statement

Hospital Clínico Universidad de Chile approved the study on health worker personnel (Protocol ID Number 1151/20 and Protocol ID Number 074-2020). Hospital Clínico Metropolitano La Florida “Dra. Eloisa Díaz I.” was included in the ethical protocols of the University of Chile as part of the COVID-19 research program of ANID grant 0752. Samples obtained from non-health worker individuals were approved by Facultad de Ciencias, Universidad de Chile (Protocol ID 2123-FCS-UCH and consent approval). Samples were collected from February 2021 to January 2022. All patients and healthy controls were required to understand the study and sign an informed consent.

### Design of study groups

We obtained blood samples from different individuals; healthcare personnel volunteers from Hospital La Florida, and adult healthy volunteers (over 18 years old). These were divided in four different groups; Group 1 to study the immune response following two Coronavac doses in healthcare personnel; Group 2 that was designed to compare Coronavac and BNT162b2 vaccination; Group 3 to analyze the homologous and heterologous booster response (6 months after Coronavac vaccination); and Group 4 to study the persistence of the humoral response after > 100 days following the homologous and heterologous booster.

This study is composed of four groups covering the period ranging from the beginning of the vaccination program in February 2021 and months after the administration of the booster doses in August 2021 (depicted as timelines in Figure 1A). Group 1 corresponds to 104 individuals belonging to the clinical staff from the Hospital Clinico Metropolitano La Florida “Dra. Eloisa Diaz”, which were among the first cohort to be vaccinated as a priority group CoronaVac vaccine. In this group of individuals, the antibody response to the first and second dose of the CoronaVac vaccine was assessed.

**Figure 1:**
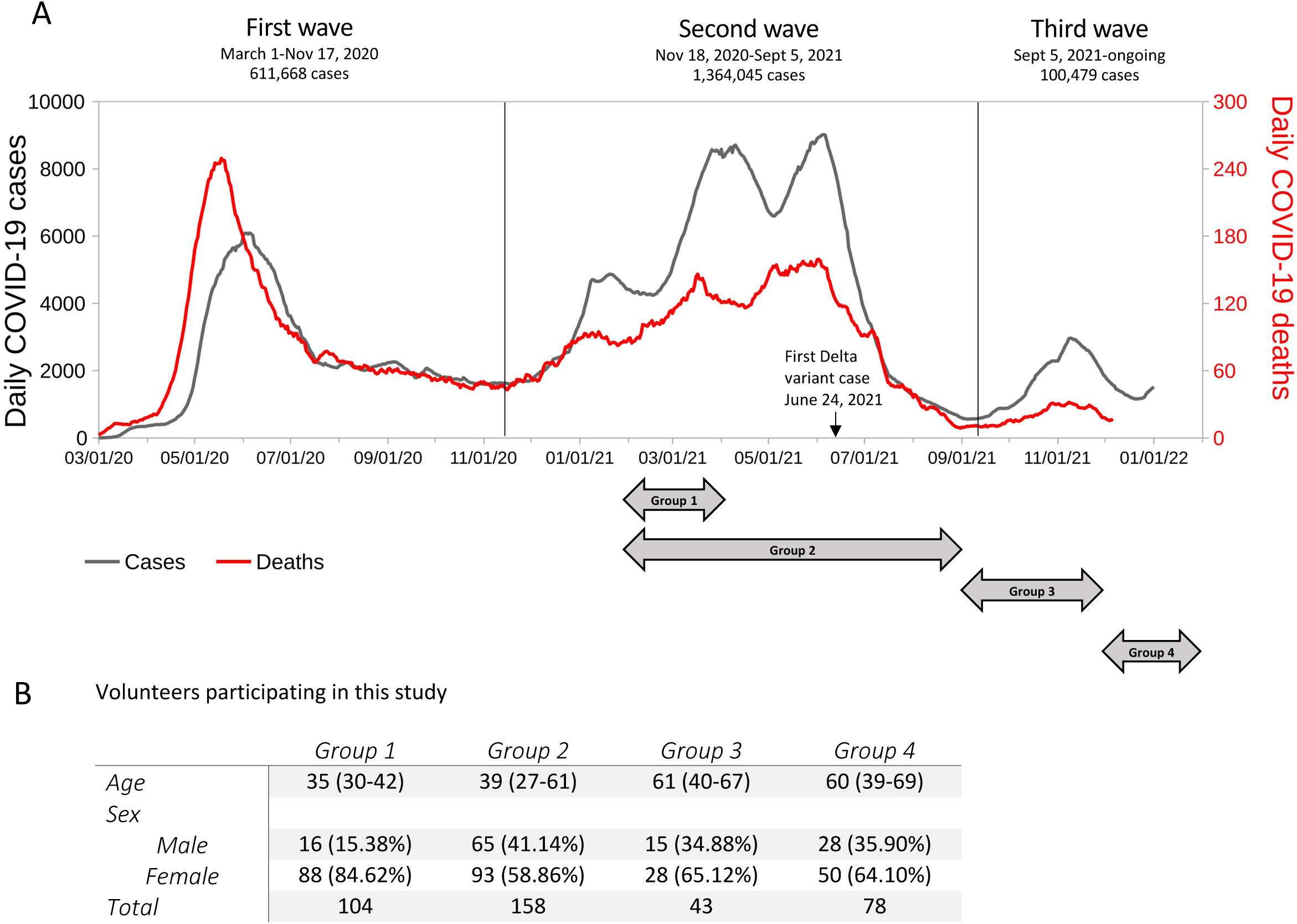
Course of the COVID-19 pandemic in Chile and details of the participants of this study. A) Chile had three waves of COVID-19. The first begun on April 2020, and ended in August 2020. The second wave was mainly caused by gamma and lambda variants and it was more extensive, beginning in November 1, 2020, and ending in September 2021 (Data extracted from MINSAL/DEIS). Finally, the delta variant entered the country on June 24^th^, 2021, and became predominant as of October 2021. The black curve depicts the daily cases of COVID-19, while the red curve represents deceased people due to COVID-19 during the period. B) Characteristics of the volunteers participating in different stages of this study. Group 1 was was composed of health care personal from HLF and was designed to test pre-immunization with CoronaVac, one month after the first and one month after the second dose. Group 2 contains samples from healthy donors recruited to compare the antibody response to CoronaVac and BNT162b2 one month after the second dose. In group 3, we studied the effect of a homologous or heterologous booster scheme in the antibody response of people vaccinated six months before with two-doses of CoronaVac. Samples were collected 20 days after the booster. In group 4, the antibody response was determined 100 days after the booster date in individuals who received two-doses of the CoronaVac vaccine originally. Values are the mean and interquartile range (IQR).

Group 2 corresponds to 158 individuals from a broad range of age vaccinated with CoronaVac and BNT162b2 vaccines. A comparison of IgG production against spike SARS-CoV-2 protein induced by the vaccines was performed, and antibody evolution was followed over time.

Group 3 corresponds to 43 individuals vaccinated with the two-dose scheme of CoronaVac that received a booster dose with either CoronaVac, BNT162b2, or the ChAdOx1 vaccine. This group determined the magnitude of the antibody response to the homologous and heterologous booster schemes 20 days after the booster. Finally, group 4 corresponds to 78 individuals vaccinated with the two-dose scheme of CoronaVac that received a booster dose with either CoronaVac, BNT162b2, or the ChAdOx1 vaccine and analyzed 100 days after the booster. Fig 1B describes the characteristics of the volunteers who participated in each stage of the study. In total, 316 individuals participated, of which 57.4% were women, and 42.6% were men. The median age of the volunteers was 38 years (Interquartile range; IQR: 30-59 years). Some individuals participated in group 2 and the longitudinal booster study. Thus, the number of samples is higher than the number of participants.

### Isolation of human blood samples

Blood samples were obtained from healthcare personnel volunteers from Hospital La Florida, and adult healthy volunteers (over 18 years old). Serum was collected after whole blood centrifugation and stored at -80ºC for further analysis.

## ELISA

The ELISA was performed as detailed (5), and adapted from the group of Kramer (6). Briefly, 96-well ELISA plates were coated overnight at 4°C with 50 μl per well of a 2 μg/ml solution of resuspended SARS-CoV-2 Spike protein (Recombinant SARS-CoV-2 S protein S1 from the original Wuhan SARS-CoV-2 virus, Biolegend 796906) on PBS. Then, the coating solution was removed, and the wells were blocked for one hour at room temperature with 150 μl of 3% skim milk prepared in PBS-0.1% Tween-20 (TPBS). After this period, 100 μl per well of serial dilutions (from 1/200 to 1/1,600) of the sera prepared in 1% skim milk in 0.1% TPBS was added and incubated for 2 hours at room temperature. The plates were washed three times, added 100 μl per well of HRP-conjugated anti-human IgG (HRP Donkey anti-human IgG Clone: Poly24109, Biolegend), and incubated for 1 hour at room temperature. The plates were washed three times, after which 50 μl of TMB substrate solution (BD Biosciences) was added per well to reveal the reaction, which was stopped by adding 50 μl per well of 1M orthophosphoric acid. Optical density at 450 nm was measured on a Molecular Devices Emax ELISA plate reader. We tested the specificity of our ELISA assay by analyzing serum samples from unvaccinated COVID19 patients at the time where Delta variant was dominant in our country. Our data confirmed that the ELISA test we performed with the S protein of the original coronavirus recognizes all the variants that have entered Chile at that time, including the Delta variant.

### Neutralization assay

#### HIV-1-based SARS-CoV-2 pseudotype production

Pseudotyped viral particles were produced by transient transfection of HEK293T cells using polyethylenimine (PEI) and plasmids pNL4.3-ΔEnv-Firefly and pCMV14–3X-Flag–SARS-CoV-2 SΔ19C (lineage A) in a 1:1 ratio as we described (7). The viral particles were diluted with 50% in fetal bovine serum (Sigma-Aldrich) and stored at -80°C. Viral stock was quantified with the HIV-1 Gag p24 Quantikine ELISA kit (R&D Systems).

Neutralizations assays were performed as we previously reported (7). Briefly, inactivated serum samples were diluted in DMEM with 10% fetal bovine serum (serial dilutions from 1:4 to 1:8748) and incubated with 5 ng of p24 HIV-1-based SARS-CoV-2 pseudotyped particles during 1h at 37°C, and 1×10^4^ HEK-ACE2 cells were added to each well. HEK293T cells incubated with the pseudotyped virus were used as a negative control. Cells were lysed 48 hours later, and firefly luciferase activity was measured using the Luciferase Assay Reagent (Promega) in a Glomax 96 Microplate luminometer (Promega). Then the percentage of neutralization for each dilution was calculated as previously described. All statistical analyses were performed using GraphPad Prism version 8.0.1 (7)

#### Quantification and statistical analysis

For the ELISA assay, the background value was established at OD 0.100, and Area Under the Curve (AUC) was calculated from serum dilutions. To obtain a correlation between AUC and antibody titers, we used estimated values of antibody titers from 212 samples, and we established a curve according to Padé’s approximation (with R2=0.9636). Differences between clinical groups were calculated using a one-way ANOVA with Freedman or Kruskal-Wallis test followed with Dunn’s multiple comparations test. Differences between the two groups were calculated using the unpaired two-tailed t-test or Mann-Whitney test. Simple linear regression was performed, and correlations were analyzed by calculating nonparametric Spearman’s correlation. Statistical analyses were performed using GraphPad Prism 9.1.0, and statistical significance was represented by *p<0.05, **p<0.01, ***p<0.001, and **p<0.0001.

## RESULTS

### The course of the humoral response to the CoronaVac vaccine

Fig 1A shows the course of the COVID-19 pandemic in Chile from March 2020 to December 2021 (MINSAL/DEIS), which illustrates three waves of COVID-19. The first wave began on April 20, 2020 and ended in August 2020. The second wave, mainly caused by gamma and lambda variants, was more extensive, beginning on November 1, 2020, and ending in September 2021 (MINSAL/DEIS). The drop in active cases began mid-June 2021 and coincided with the drop in cases throughout South America (MINSAL / DEIS, our world in data). Finally, the delta variant entered the country on June 24th, 2021, and became predominant as of October of this year causing the third wave. However, this variant was notably less infectious in Chile than in European countries. Additionally, in the last days of November, the entry of the first case of the omicron variant was reported.

This study is composed of four groups of individuals that were analyzed across entire the vaccination and booster programs in 2021, starting February 2021 and ending in January 2022 (depicted as timelines in Figure 1A). The number of individuals and additional details of the study are found in Fig 1B.

### Serological analysis of CoronaVac before immunization, and post-first and -second dose

To evaluate the effect of the CoronaVac vaccine on antibody titers in individuals potentially exposed to the SARS-CoV-2 virus, we first focused our study on clinical staff from group 1, who treated COVID-19 patients in the first wave of the disease in Chile. We analyzed the serum of these individuals by ELISA to detect IgG antibodies directed against the Spike (S) protein of the SARS-CoV-2 virus. This test was developed with samples from hospitalized COVID-19 patients as positive controls (13 samples) and pre-pandemic or negative samples (54 samples) for negative controls (5) and developed as reported (6). Sera were diluted serially from 1/200 to 1/1,600, and the area under the curb (AUC) was determined. These values were equivalent to the antibody titer (see Methods). We established the negative limit of the test (AUC = 70 ± 51) from the analysis of 54 samples from people who had no history of COVID-19. We considered AUC values between 120 and 300 as a weak response in the ELISA test. In contrast, an AUC of around 300 corresponds approximately to an antibody titer of 1/1,000. To analyze the SARS-CoV-2 antibody response course in this group, we analyzed the antibody response in three-time points. The first serum sample was obtained 1-3 days before the first dose of the vaccine (referred to as ‘Preimmune’, Fig 2); the second sample was obtained 1-3 days before the second immunization (referred to as ‘First dose + 30d’, Fig 2), and the third sample was collected one month after the second dose (referred to as ‘Second dose +30d’, Fig 2). Regarding previous SARS-CoV-2 infection, the individuals who participated in this study were laboratory staff, and primary clinical caregivers in contact with COVID-19 patients. Many individuals in this group reported not knowing whether they had been exposed to SARS-CoV-2 since they could have experienced the asymptomatic disease.

**Figure 2:**
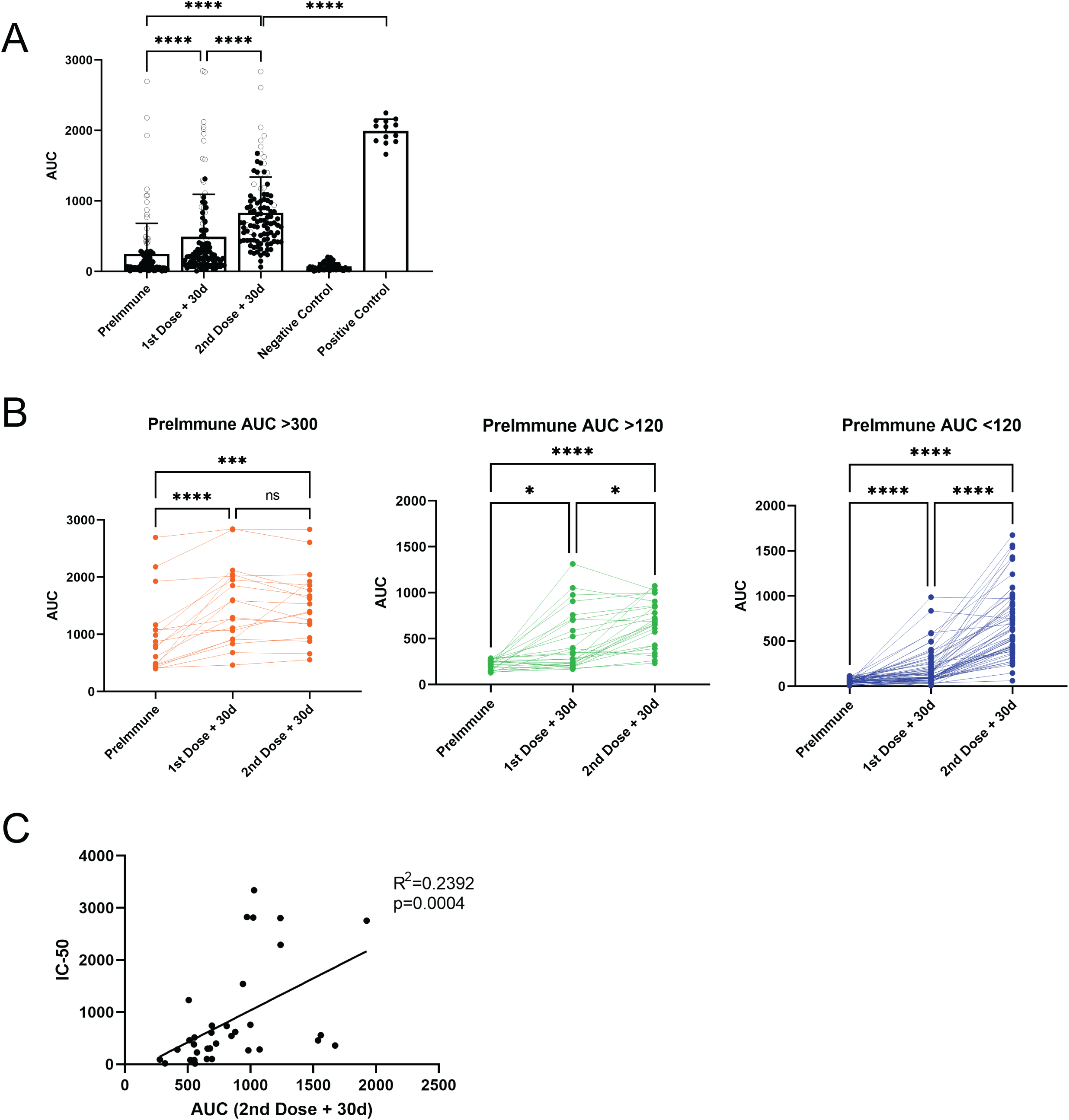
Serological analysis of CoronaVac before immunization, and post-first and -second dose. Correlation with neutralization antibodies. Health Care services volunteers received a complete CoronaVac vaccination scheme. Serum samples were collected as indicated: PreImmune, prior to the first dose, first dose + 30d, 30 days after the first dose, second dose + 30d, 30 days after the second dose. A) and B) Serum reactivity to SARS-Cov-2 S protein was expressed as 21imp under the curve (AUC) obtained from four serial dilutions from 1/200 to 1/1,600 for each 21imple. A) Data from 104 volunteers are shown before vaccination, 30 days after the first, and 30 days after the second dose. The gray circles show the values of people who contracted the disease before vaccination. Black circles are from the other 86 samples being negative or weakly positive. Negative controls were obtained from 54 pre-pandemic or COVID-19 negative samples and 13 positive controls from COVID-positive patient samples. Significance was assessed by nonparametric Friedman test with Dunn’s multiple comparisons test, and comparison between positive control and second dose +30d was determined with Mann-Whitney test. B) Data shown in A) were disaggregated into three groups 21imple21n AUC values before vaccination: 18 Individuals with an AUC> 300 (positive) are shown in orange, 26 with an AUC> 120 (weakly positive) in 21impl, and 60 individuals negative for SARS-CoV-2 prior to vaccination in blue. In each case, significance was assessed by the nonparametric Friedman test with Dunn’s multiple comparisons test. **C** Neutralization assay using CELLS DEL ENSAYO. C) SerumC neutralization capacity in vaccinated participants 30 days after the second dose was correlated with correspondent AUC. Significance was assessed with Spearman’s 21imp correlation, and simple linear regression determined the R2 value. Each dot represents a single serum 21imple. ****p<.0001 ***p<.001 *p<.05.

Of a total of 104 people tested, 18 had high antibody titers (AUC> 300) before being vaccinated, suggesting that these individuals were infected with SARS-CoV-2 in the first pandemic wave (Fig. 2A, empty circles). Of these 18 individuals, only two did not improve antibody titers with vaccination (Fig. 2B orange lines). For the remaining 16 individuals (orange lines), the first dose of the vaccine led to an increase in anti-S IgG production. Interestingly, there were no statistical differences when comparing the level of antibodies induced by the first and the second dose of the vaccine (Fig. 2B, orange circles).

For individuals who initially had an AUC>120 (weak positive reaction, Fig 2B green circles), the first dose showed an increase in the level of anti-S IgG. Although significant, there was a mild difference between the first and the second doses. Interestingly, the group who initially had an AUC<120 (negative reaction, Fig 2B blue circles) showed remarkable differences between the first and the second dose of the vaccine. Of the total 104 people, only one person remained unresponsive to the two doses of the vaccine. As such, we conclude that the two-dose vaccination scheme with CoronaVac induces a good antibody response against SARS-CoV2, which is particularly noticed in individuals who have not been previously exposed to the virus.

Next, the amount of neutralizing antibodies from 34 samples obtained one month after the second dose was determined. The results show a significant positive correlation between the AUC values and the IC-50 of neutralizing antibodies (Fig. 2C). These results demonstrate that the CoronaVac vaccine induces the production of neutralizing antibodies. Furthermore, this data suggests that high titers of total antibodies should represent a greater probability of having neutralizing antibodies against the virus.

### Comparison of the humoral immune responses produced by the CoronaVac and BNT162b2 vaccines

The first reports of CoronaVac vaccine immunogenicity were performed in older adults (over 55 years old) (8) since these individuals were among the priority groups for vaccination. In May 2021, individuals under 55 years old began to be vaccinated with BNT162b2 or CoronaVac depending on the availability of the vaccine in Chile. This allowed us to analyze the antibody response 30 to 45 days after the second dose to compare the humoral response elicited by both vaccines. We studied 44 and 20 individuals vaccinated with CoronaVac and BNT162b2 (Pfizer-BioNTech) vaccine, respectively (group 2). Figure 3A shows a comparison of the data from both vaccines in individuals raging from 18-87 years old (IQR: 27-61 years). We observed that the BNT162b2 vaccine induces significantly higher antibody production than the CoronaVac vaccine (2060 ± 361 for BNT162b2 and 1041 ± 520 for CoronaVac). Given that people vaccinated with CoronaVac were mainly older than 55 years in Chile and those vaccinated with BNT162b2 were people between 18 and 54 years old, we compared and plotted antibody production according to the age of the individuals and the type of vaccine they received. Figure 3B shows a significant negative correlation (p = 0.032, black circles) for antibody production with increasing age for the CoronaVac vaccine. In contrast, a similar (but not statistically significant, green circles) trend is shown for the BNT162b2 vaccine. These results show that the BNT162b2 vaccine induces twice the amount of IgG against SARS-CoV-2 S protein compared to CoronaVac, independent of the age of the individuals.

**Figure 3.**
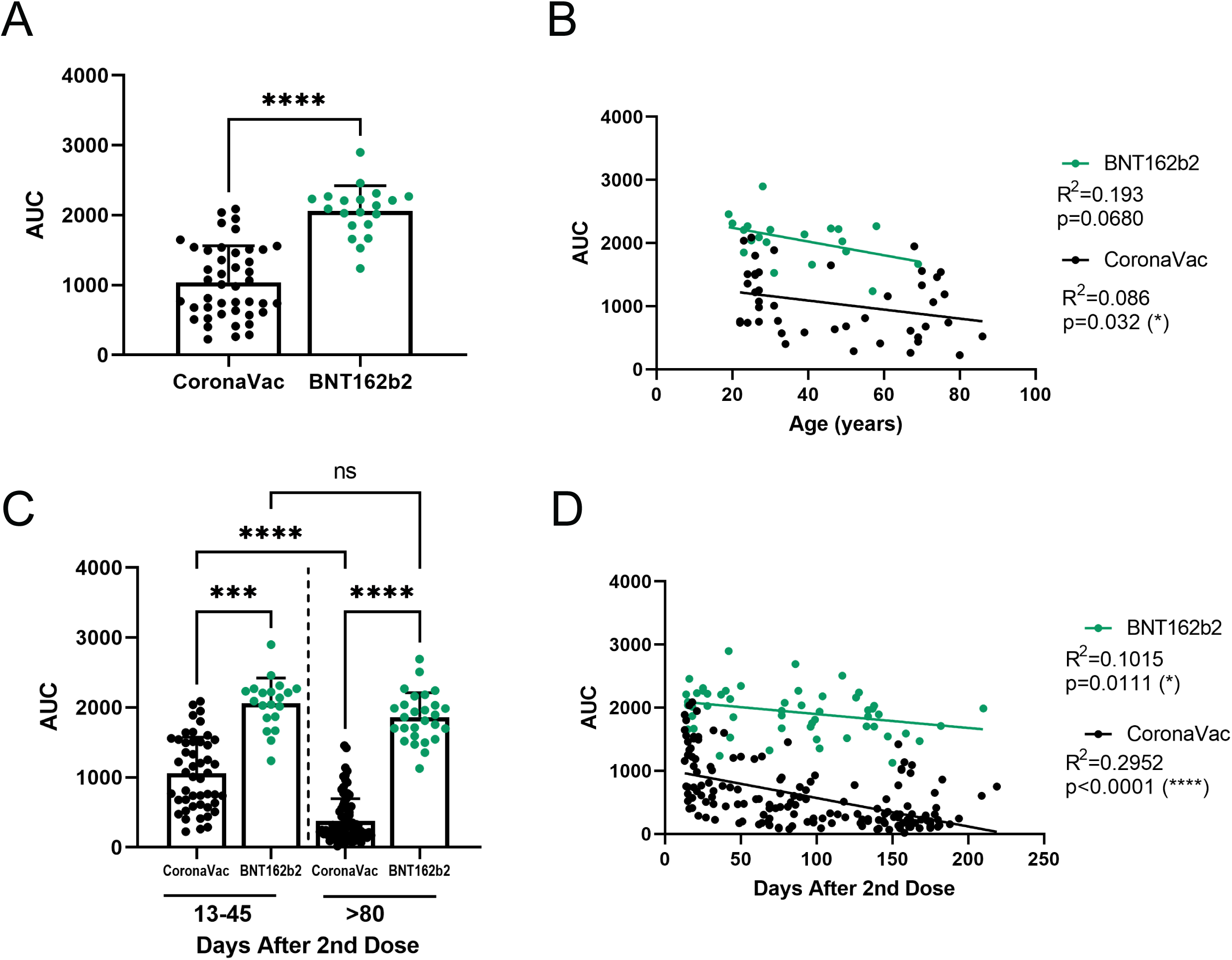
Comparison of antibody response to CoronaVac and BNT162b2 vaccines over time and range of age. Healthy participants received a complete vaccination scheme, and serum samples were collected after the second dose at the indicated time. A) and B) Comparations between the antibody titers of samples obtained between 30 to 45 days after the second dose. Forty-four individuals vaccinated with CoronaVac and 20 with BNT162b2 (green) A) Direct comparison between the antibody titers against SARS-CoV-2 S protein of CoronaVac (black) or BNT162b2 (green) vaccine. A nonparametric Mann-Whitney test assessed significance. B) Correlation between age and antibody titers of individuals vaccinated with CoronaVac and BNT162b from the serum of first 45 days. Significance was assessed by Spearman correlation with significant value for CoronaVac, and simple linear regression determined R2 value. Each dot represents a single serum sample. C) and D) Antibody titers from 138 samples collected more than 80 days after the second dose. C) Samples from 13 to 45 days were compared to samples from more than 80 days from CoronaVac (black) or BNT162b2 (green) vaccine scheme; nonparametric Kruskal-Wallis assessed significance with Dunn’s multiple comparisons test. Each dot represents a single serum sample. D) Correlation between age and antibody titers of individuals vaccinated with CoronaVac and BNT162b along time. Significance was assessed by Spearman correlation with significant value for both vaccines and simple linear regression determined R2 value. Each dot represents a serum sample. ****p<.0001 ***p<.001 *p<.05.

### Overtime evolution of the humoral response to CoronaVac and BNT162b2 vaccines

So far, we have demonstrated the presence of neutralizing antibodies in a significant number of individuals immunized with CoronaVac and demonstrated a positive correlation between the amount of IgG against SARS-CoV-2 S antibodies and the production of neutralizing antibodies (Fig. 2C). Moreover, we showed that the BNT162b2 vaccine produces higher levels of antibodies in vaccinated people than those elicited by the CoronaVac vaccine (Fig. 3A). Therefore, we sought to determine how antibody levels vary with these two vaccines over time. For this purpose, we analyzed samples taken 15 to 200 days after the second dose of CoronaVac or BNT162b2 vaccines. One hundred and fifty-nine samples from individuals vaccinated with CoronaVac and 53 samples from individuals vaccinated with BNT162b2 were analyzed. Fig. 3D shows a significant negative correlation for each of these vaccines (CoronaVac p <0.0001; BNT162b2 p = 0.0111). The curve slope allows us to infer that around 200 days after the second dose of the CoronaVac vaccine, most individuals vaccinated will present low antibody titers against the SARS-CoV-2 S protein. In contrast, in individuals vaccinated with the BNT162b2 vaccine, antibodies slightly decrease in most individuals, agreeing with data from the literature (2).

We then disaggregated the data to visualize the results (Fig. 3C). Comparing the data obtained 13 to 45 days or beyond 80 days after the second dose from both vaccines, we observed a significant loss of antibodies beyond 80 days after the second dose of the CoronaVac vaccine (1,057±519 vs. 378±318) compared to the BNT162b2 vaccine (2,060±361 vs. 1,861±351). These data suggest that the BNT162b2 vaccine is more efficient in inducing and maintaining the production of antibodies against the SARS-CoV-2 virus S protein.

### Analysis of the antibody response of individuals receiving homologous or heterologous booster dose schemes

A total of 44 individuals who were vaccinated with two doses of CoronaVac received, around 180 days after the second dose, a booster dose with the ChAdOx1 vaccine (19 individuals), BNT162b2 vaccine (19 individuals), or CoronaVac vaccine (5 individuals) (timeline scheme depicted in Fig. 4A). Data illustrated in Fig. 4B show that regardless of the type of vaccine used for the booster dose, all individuals significantly enhanced IgG production against the Sars-CoV-2 S Protein. Values range from 268±218 before the boost to 2,245±581 considering any booster, meaning an 8,37-fold change average. However, when we separated the data based on the type of booster vaccine, we observed that the CoronaVac booster vaccine-induced antibody production, which was noticeable but milder (fold induction: 9.8x) than the antibody production induced by the ChAdOx1 vaccine booster (fold induction: 12.4x) or the BNT162b2 vaccine booster (fold induction: 11.2x). These results demonstrate that the CoronaVac vaccine combined with a booster from CoronaVac or any other vaccine enables memory immune response to be activated, in agreement with recent data (9). These authors showed that a booster with CoronaVac vaccine eight months after the second dose increased neutralizing antibodies against the original virus SARS-CoV-2. However, it is noteworthy to mention that the antibody response induced by the third dose of the CoronaVac vaccine is lower than the two other boosters.

**Figure 4.**
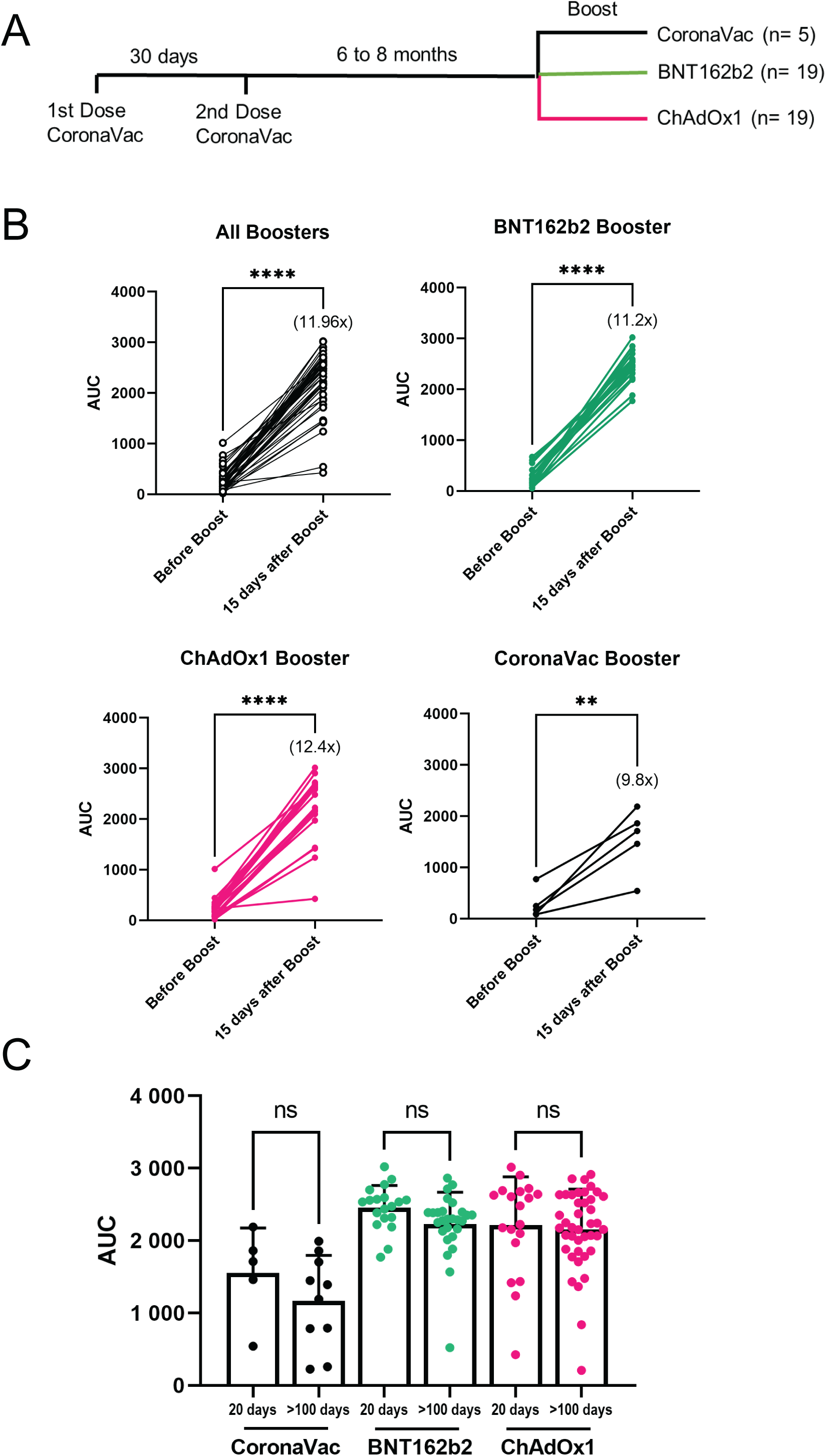
Antibody titers of homologous and heterologous boosters of individuals previously vaccinated with two doses of CoronaVac. Participants received a complete CoronaVac vaccination scheme and booster after 6 to 8 months with BNT162b2, ChAdOx1, or CoronaVac vaccine. Serum samples were collected as indicated: Before Boost and 20 days after Boost to evaluate the change of antibody titer. A) Schema of participant’s immunizations. B) Antibody titer comparison before and 15 days post booster immunization from BNT162b2 (19 individuals) ChAdOx1 (19 individuals), and CoronaVac (5 individuals). C) Antibody titer comparison 100 days post booster immunization (Median 128 days; IQR: 119-135 days) from BNT162b2 (27 individuals) ChAdOx1 (41 individuals), and CoronaVac (10 individuals). Significance was assessed by paired parametric t-test (‘BNT162b2’ and ‘CoronaVac’) or paired nonparametric Willcoxon test (‘All Boosters’ and ‘ChAdOx1’). Each dot represents a serum sample. ****p<.0001 **p<.01.

To obtain insights on the extension of the antibody response induced by the homologous and heterologous booster regimes, we measured anti-SARS-CoV-2 S antibodies in 78 individuals 100 days after the booster dose (Median 128 days; IQR: 119-135 days) (Fig. 4C). This analysis revealed that the homologous booster with CoronaVac showed a trend towards a decline in antibody production, which did not reach statistical significance (Fig. 4C). However, the antibody response elicited by heterologous boosters with BNT162b2 and ChAdOx1 vaccines remained higher than the homologous scheme and did not show noticeable signs of immunological waning during the period. Overall, our results suggest that a heterologous booster scheme using CoronaVac as the basal vaccine with a booster from ChAdOx1 (AstraZeneca) vaccine or BNT162b2 (Pfizer) vaccine, re-activate immune memory and elicits a potent and persistent immune response at least over a 3-month period.

## DISCUSSION

This work reports the dynamics of anti-S IgG after SARS-CoV2 vaccination with CoronaVac, a vaccine used globally (10), a comparison with an mRNA vaccine over time, and an assessment of homologous and heterologous booster schemes in Chile using CoronaVac as the basal vaccine. The groups analyzed in this study span the entire vaccination program in Chile, from the beginning of the vaccination schedule with priority groups, to the implementation of booster schemes in late 2021.

Our data indicate that in individuals not exposed to SARS-CoV-2, a two-dose vaccination scheme with CoronaVac induces a noticeable antibody response against SARS-CoV-2, in agreement with additional reports (9). Furthermore, there is a positive correlation between the production of neutralizing antibodies and those detected by ELISA (AUC). When comparing CoronaVac and BNT162b2 vaccines, we found that the BNT162b2 vaccine is more efficient in inducing and maintaining the production of antibodies against the SARS-CoV-2 virus S protein independent of the age of the individuals. Moreover, we evaluated three different booster schemes in people previously vaccinated with CoronaVac. We found that a homologous booster with CoronaVac or heterologous boosters with ChAdOx1 (AstraZeneca) vaccine or BNT162b2 (Pfizer) vaccine can elicit a humoral immune response against the ancestral strain of the virus. However, our data strongly indicates that heterologous booster regimes greatly potentiate antibody responses compared to a homologous regime. As such, our findings may have relevant implications for the large number of countries currently administering a two-dose scheme of CoronaVac.

Concerning the booster schemes, administration of a homologous booster scheme of CoronaVac has been demonstrated to be immunogenic and safe in a double-blind, randomized, placebo-controlled phase-2 clinical trial (9). In this context, the homologous and heterologous booster schemes analyzed in this work re-activated anti-S IgG production in individuals previously vaccinated with the two-dose scheme of CoronaVac. Analysis over a more extended period of time (more than 100 days) revealed that heterologous booster schemes are capable of inducing an elevated and long-lasting antibody response compared to two-doses plus a booster of CoronaVac. Thus, these data suggest that the use of heterologous instead of homologous booster regimes may allow to space the subsequent booster doses to achieve long-lasting humoral response and protection against COVID19. These findings also provide evidence that will allow to prioritize the subsequent booster doses in individuals that have lost optimal anti-SARS-CoV-2 antibodies, such as those with the homologous regime.

It remains to be observed if these heterologous regimes potentiate an immune response that could provide protection (or partial protection) against novel variants. In this context, many questions remain to be addressed. For instance, although we provide data of over 3 months after the booster, it is unclear how long the protection mediated these booster schemes will last or if these strategies will efficiently protect against novel variants such as delta and the recently described omicron (11). In this regard, a very recent study of a heterologous booster scheme based on CoronaVac + BNT162b2 in the Dominican Republic showed a reduced antibody response towards the Omicron variant (12). One distinction between that study and the data presented here relates to the timing between the second dose and the booster, which in Chile was implemented after a six-month interval, whereas in the Dominican Republic study, the heterologous booster scheme was implemented after four weeks (12). As such, the immune response elicited under two different time schemes may differ in terms of the magnitude of antibody production. Thus, future work combined with clinical studies are required to determine the optimal time between vaccine and booster administration. Along these lines, the study of Zeng et al demonstrates that extending the interval of eight months between the second and the homologous booster dose with CoronaVac greatly increases antibody production (9). Interestingly, our study also reports potent responses with the heterologous booster scheme with the ChAdOx1 vaccine, requiring further assessment. In addition, our work is also in line with a very recent report showing that heterologous booster regimes are superior to homologous booster schemes based on the CoronaVac vaccine in a Brazil study (13).

One limitation of our study is that we assessed antibody production against the spike protein of SARS-CoV2 but a relevant response mediating long-lasting immunity could also be carried out by T cells, which are not analyzed in this work. However, a recent study with 15 volunteers with no suspected history of COVID-19, vaccinated with two doses of CoronaVac showed humoral and cellular immune response 28d after the second dose (14).

As such, it is possible that a heterologous booster scheme based on CoronaVac as the basal vaccine could lead to potent immunity, based on the diversity of viral antigens provided by an inactivated virus formulation, followed by a booster with mRNA or adenoviral vector vaccines, which trigger a superior degree of immunogenicity. The long-term immunological effects related to protection against SARS-CoV-2’ variants of concerns and variants of interests induced by heterologous booster strategies should be determined with high priority in order to shed light on the future management of the pandemic across the globe.

## Data Availability

All data produced in the present work are contained in the manuscript.

## Acknowledgments

We thank all the volunteers who collaborated in this work, particularly the Clinical Laboratory of the La Florida Hospital staff, the staff who work at the Juan Gomez Millas Campus, and many other volunteer’s collaborating with the study. Thanks to the 2021 generation Biotechnology Engineering students for their interest in participating in a real-world experiment.

## Author contributions

Conceptualization: MRB/FO/ML/LC; Data Curation: MRB, LV, VS; Formal Analysis: LV/NV/MLA; Funding Acquisition: MRB/FO/ML/RSR/FV; Investigation: MRB/FO/ML; Methodology: LV/NV/MLA; Project Administration: MRB/LV/VS; Resources: MRB/FO/ML/LC/CB/LV; Supervision: MRB; Validation: LV/NV/MRB/FO/RSR; Visualization: NV/DS/RG; Writing-Original Draft Preparation: MRB/FO; Writing-Review & Editing: MRB/FO/ML/DS.

